# Advancing Public Health Through Building Physician Capacity for Salt Reduction and Low Sodium Substitutes

**DOI:** 10.64898/2026.06.18.26355944

**Authors:** Ravichandran Rajan, S. Kerenhappuch Susan, Manika Sharma, Umesh B. Khanna, Vijay Kher, Bharat Shah, Edwin Fernando, K. Sampathkumar, S.N. Narasingan, N. Murugesan, R. Krishna Kumar, Zamrrud Patel, Sundar Ramachandran

**Author notes:** Corresponding Author Dr. Ravichandran Rajan Sapiens Health Foundation Rams Flats, 95/42, Second Main Rd, Gandhi Nagar, Adyar, Chennai, Tamil Nadu 600020.

## Abstract

Excessive sodium intake is a major contributor to hypertension, cardiovascular disease, and other non-communicable diseases (NCDs), posing a persistent public health challenge. Physicians, as trusted health advisors, are central to promoting salt reduction strategies in routine clinical practice.

This study describes a physician capacity-building initiative conducted by Sapiens Health Foundation in three metropolitan cities of India: Chennai, Mumbai, and Delhi, between June 2024-February 2025. The workshops aimed to strengthen physician knowledge and counselling skills regarding dietary salt reduction and the use of low-sodium salt substitutes (LSS). Each workshop featured expert presentations, interactive discussions, and evidence-based educational resources. Evaluation methods differed by location: Chennai and Mumbai used structured feedback forms, while Delhi implemented pre- and post-training assessments to measure knowledge gains.

Findings showed that while many physicians were aware of the WHO’s recommended salt intake, gaps remained in areas such as hidden salt sources, sodium-to-salt conversions, low sodium salt (LSS) substitutes and appropriate use of LSS in clinical practice. Post-training results indicated improved knowledge across all cities, with statistically measurable gains in Delhi.

This initiative demonstrated that structured workshops can enhance physician capacity in salt reduction and dietary counselling and encourage them to act as advocates within clinical and community settings. Integrating such training into medical education and aligning it with national public health strategies can help reduce salt consumption at the population level. Wider adoption of this model, supported by digital learning tools, could improve health outcomes and reduce the burden of NCDs in India.

## Introduction

Reducing excessive sodium intake is essential to public health improvement, especially in combating non-communicable diseases (NCDs) such as hypertension and cardiovascular disease, which remain leading contributors to global morbidity and mortality [1]. Given their trusted role and direct access to patients, physicians are central to advancing salt reduction strategies [2,3]. Their involvement bridges the gap between public health recommendations and individual dietary behavior, making them powerful agents for both prevention and policy change [4].

Through routine clinical encounters, physicians have a unique opportunity to educate patients about the risks of high salt intake and promote sustainable dietary changes. Evidence shows that many primary healthcare providers already recognize the importance of sodium reduction and frequently advise patients to adopt lower-salt diets [5]. In the United States, a considerable proportion of adult’s report receiving such advice directly from their healthcare providers [2]. This heightens the importance of enhancing physicians’ capacity to effectively counsel patients on salt reduction and the broader role they play in shaping public health messaging for India and other countries.

While building physician knowledge and skills is a key step in reducing NCD risk, improving treatment outcomes at scale also requires broader systemic changes. Physician training must be integrated within a coordinated public health framework that includes policy, community engagement, and health system support [6]. This highlights the need for a multi-component approach where clinical practice improvements are aligned with larger-scale interventions.

To achieve this, physician capacity building must be comprehensive and multi-level targeting individuals, institutions, and policy frameworks simultaneously. Training programs should cover core content such as salt-related health risks, clinical counselling techniques, and population-level interventions, information on the use of LSS and counselling messages and methods.

Expanding physician capacity further requires engagement with multi-sectoral stakeholders. Collaboration with policymakers, education departments, and food regulation authorities helps drive policy-level changes, such as reformulation of processed foods, marketing restrictions, taxation, and mandatory front-of-pack labelling including for high salt products. Involving physicians from both the public and private sector as champions and advocates could help highlight the need for such policy and programmatic changes.

By adopting an integrated, multi-level approach to physician capacity building, countries can amplify their salt reduction efforts and contribute meaningfully to reducing the burden of NCDs. Sustained investment in these strategies is essential to achieving long-term improvements in population health outcomes. Thus, workshops were conducted to improve physicians’ knowledge and practical capacity in salt reduction and the use of low-sodium salt substitutes.

### Program Implementation

The physician capacity-building program on salt reduction and the use of low-sodium salt substitutes was conducted in three metropolitan cities Chennai, Mumbai, and Delhi, between June 2024 and February 2025. The initiative was organized by Sapiens Health Foundation, in collaboration with the Directorate of Public Health and Preventive Medicine, Government of Tamil Nadu, and Resolve to Save Lives (RTSL), a global public health organization.

Each workshop was structured as half day, in-person training session aimed at general physicians, internists, and primary care providers. The sessions included a combination of expert-led presentations, interactive discussions, and distribution of educational resources. Presentations addressed key topics such as the physiological effects of excessive salt intake on cardiovascular and kidney health, the role of modifiable risk factors in non-communicable diseases (NCDs), the use and benefits of low-sodium salt substitutes, and strategies for salt reduction and counselling & public advocacy.

A standardized training manual and visual toolkit, comprising evidence-based guidelines, infographics, and patient education posters, were provided to all participants. Low-salt recipe suggestions and regional dietary examples were included to emphasise on balancing taste with health promotion. The content and speakers were carefully selected to reflect national priorities in NCD prevention and were supported by institutional leaders and public health authorities in each city.

The workshops also fostered discussion on public policy, including the need for mandatory front-of-pack warning labels, reformulation incentives for the food industry, and taxation of high fat, sugar and salt packaged foods. A particular emphasis was placed on capacity building regarding LSS as a practical sodium reduction intervention. Considering the high burden of hypertension and cardiovascular disease in India, physicians were informed on the role of LSS in salt reduction strategies, highlighting its potential to reduce population-level sodium intake while maintaining taste and dietary adherence. These deliberations reinforced the importance of equipping physicians with the knowledge and skills on latest scientific evidence and practical applicability of sodium reduction interventions including LSS. Importantly, the workshops helped initiate the development of a network of “champion physicians” who can promote salt reduction within their practice settings and wider communities. These champions are expected to play an important role in maintaining progress, guiding colleagues, and contributing to future public health programs on dietary sodium reduction.

Engagement with institutional leaders (IIT Madras Director, Public Health Department officials) and state and national stakeholders including the Directorate of Public Health (Tamil Nadu), Brihanmumbai Municipal Corporation (BMC), NITI Aayog, and the Directorate General of Health Services (DGHS), helped elevate the initiative as a scalable model for physician engagement in preventive health. The participation of these stakeholders was significant, as they represent national and state-level policy and implementation bodies. Their involvement helps link evidence-based nutrition strategies with policy and programs for large-scale impact across diverse health systems in India.

Participant evaluation methods differed across locations. In Chennai and Mumbai, structured response e-forms were administered prior to the workshops to collect data on physician awareness, perceived confidence in patient communication, and willingness to adopt and recommend low-sodium alternatives in clinical practice. Following the workshops, participants completed a knowledge-assessment questionnaire to evaluate changes in understanding. In Delhi workshop, a revised and more structure approach was implemented with pre- and post-training knowledge assessment questionnaires, to systematically measure knowledge improvement across key domains, including salt intake, health outcomes, and the use of low-sodium salt substitutes. Participation in these assessments was voluntary and not mandatory, though all physicians were encouraged to respond to ensure a more comprehensive understanding of learning outcomes. However, the data presented does not reflect all workshop participants. While the overall attendance was higher (Table 1), only a portion of physicians completed the assessments, and the findings are therefore limited to this subset of respondents.

**Table 1.**
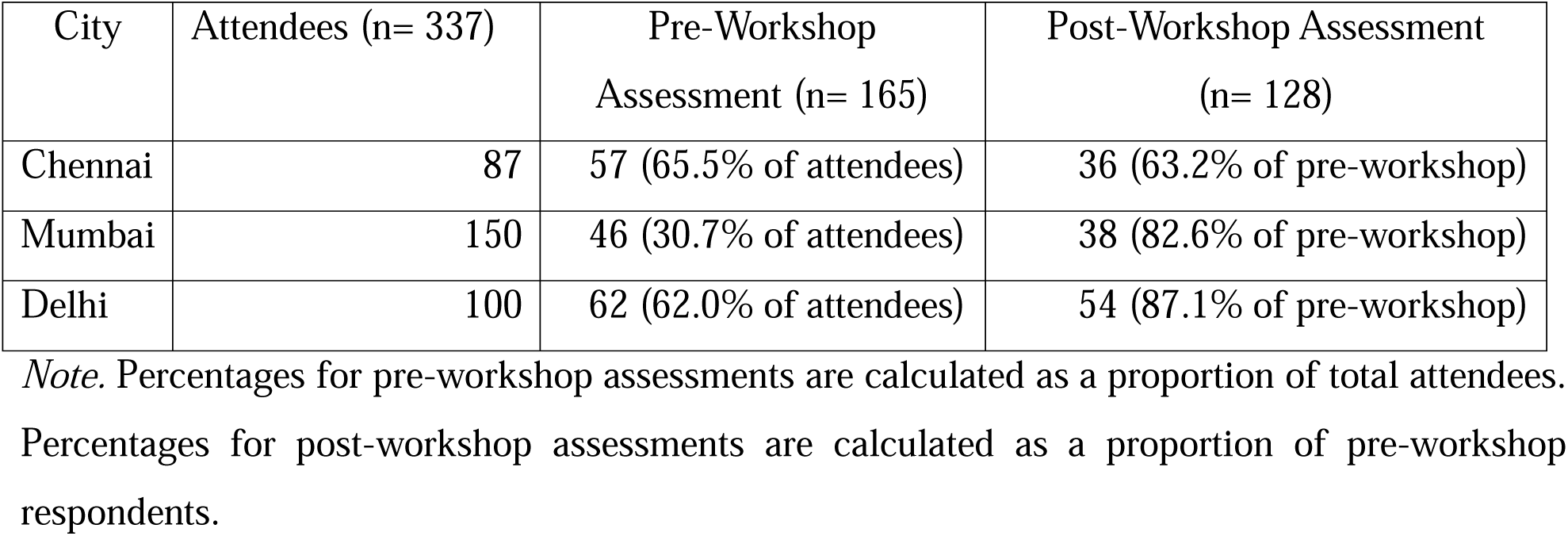
Participation in Pre- and Post-Workshop Assessments Across Three Cities.

Overall, the program successfully positioned physicians as frontline champions in India’s salt reduction efforts and laid the foundation for potential government-aligned scale-up for both the public and private sectors.

## Statistical Analysis

Data obtained from the physician assessments were entered into Microsoft Excel and analysed using IBM SPSS Statistics version 23.0 (IBM Corp., Armonk, NY, USA). Descriptive statistics were used to summarize participant characteristics and responses. Categorical variables were presented as frequencies and percentages, while continuous variables were expressed as mean ± standard deviation (SD).

For Chennai and Mumbai, post-workshop knowledge scores were compared between the two cities using an independent samples t-test. In Delhi, mean knowledge scores obtained before and after the workshop were compared using a paired samples t-test. A two-sided p-value <0.05 was considered statistically significant.

Knowledge assessment results for individual items were summarized as the number and percentage of correct and incorrect responses. Due to variations in assessment methods and response rates across the three cities, comparisons between locations were interpreted descriptively and with caution.

### Workshop Outcomes

Although the total number of attendees was higher in all three cities, only a subset of physicians completed the assessments. Mumbai and Delhi had larger numbers of total attendees. Chennai and Delhi achieved relatively higher participation in pre-workshop assessments (65.5% and 62.0%, respectively), while Mumbai recorded a lower rate (30.7%). Post-workshop participation declined in all three cities, although Delhi showed stronger retention (87.1%) between pre- and post-workshop assessments (Table 1). This indicates variability in response rates across locations, which may limit the comparability of findings and suggests the need for standardized and more engaging assessment approaches in future programs.

Overall awareness about the recommended daily salt intake and complications beyond hypertension due to excessive salt was relatively high among respondents from both Chennai and Mumbai. Mumbai participants showed notably higher awareness levels than Chennai participants for most issues, especially regarding “hidden salt” and “composition of low salt substitute.” However, substantial knowledge gaps were evident across both cities in areas like permitted salt intake for children and sodium-to-salt conversions (Table 2).

**Table 2.** Awareness of Salt-Related Knowledge Among Physicians in Chennai and Mumbai at baseline (n=103)

| Items | Place |  |  |  |  |  |
| --- | --- | --- | --- | --- | --- | --- |
|  | Chennai (n=57) |  | Mumbai (n=46) |  | Total (n=103) |  |
|  | Yes | No | Yes | No | Yes | No |
| WHO's recommended salt intake per day | 38 (66.7%) | 19 (33.3%) | 43 (93.5%) | 3 (6.5%) | 81 (78.6%) | 22 (21.4%) |
| Meaning of hidden salt | 23 (40.4%) | 34 (59.6%) | 39 (84.8%) | 7 (15.2%) | 62 (60.2%) | 41 (39.8%) |
| Can normotensive individuals consume high salt? | 2 (3.5%) | 55 (96.5%) | 7 (15.2%) | 39 (84.8%) | 9 (8.7%) | 94 (91.3%) |
| Complications of high salt intake beyond hypertension | 49 (86.0%) | 8 (14.0%) | 43 (93.5%) | 3 (6.5%) | 92 (89.3%) | 11 (10.7%) |
| Recommended salt intake for children | 21 (36.8%) | 36 (63.2%) | 29 (63.0%) | 17 (37.0%) | 50 (48.5%) | 53 (51.5%) |
| Sodium content in 1g common salt | 22 (38.6%) | 35 (61.4%) | 28 (60.9%) | 18 (39.1%) | 50 (48.5%) | 53 (51.5%) |
| Composition of low-sodium salt substitute | 16 (28.1%) | 41 (71.9%) | 28 (60.9%) | 18 (39.1%) | 44 (42.7%) | 59 (57.3%) |

Post-workshop results from Chennai and Mumbai showed significant improvement in physicians’ understanding of core topics related to salt reduction, particularly on WHO-recommended limits and sources of dietary salt. Correct responses were high across both cities, especially in questions related to salt substitutes and their safe use. However, complex clinical aspects, like hyperkalaemia management and dietary assessment methods, had lower scores, suggesting a need for enhanced focus on clinical application and risk communication in future trainings (Table 3).

**Table 3.**
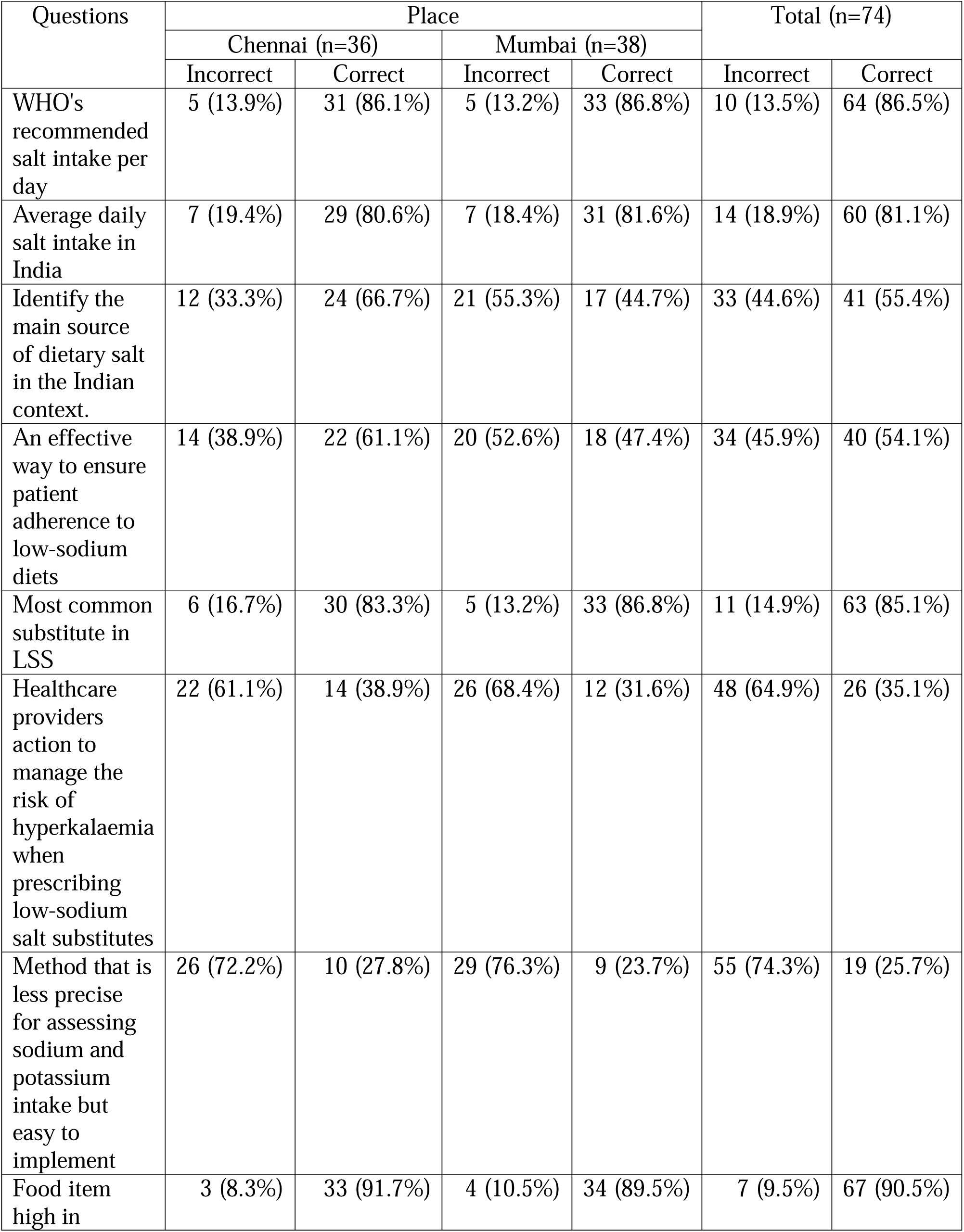
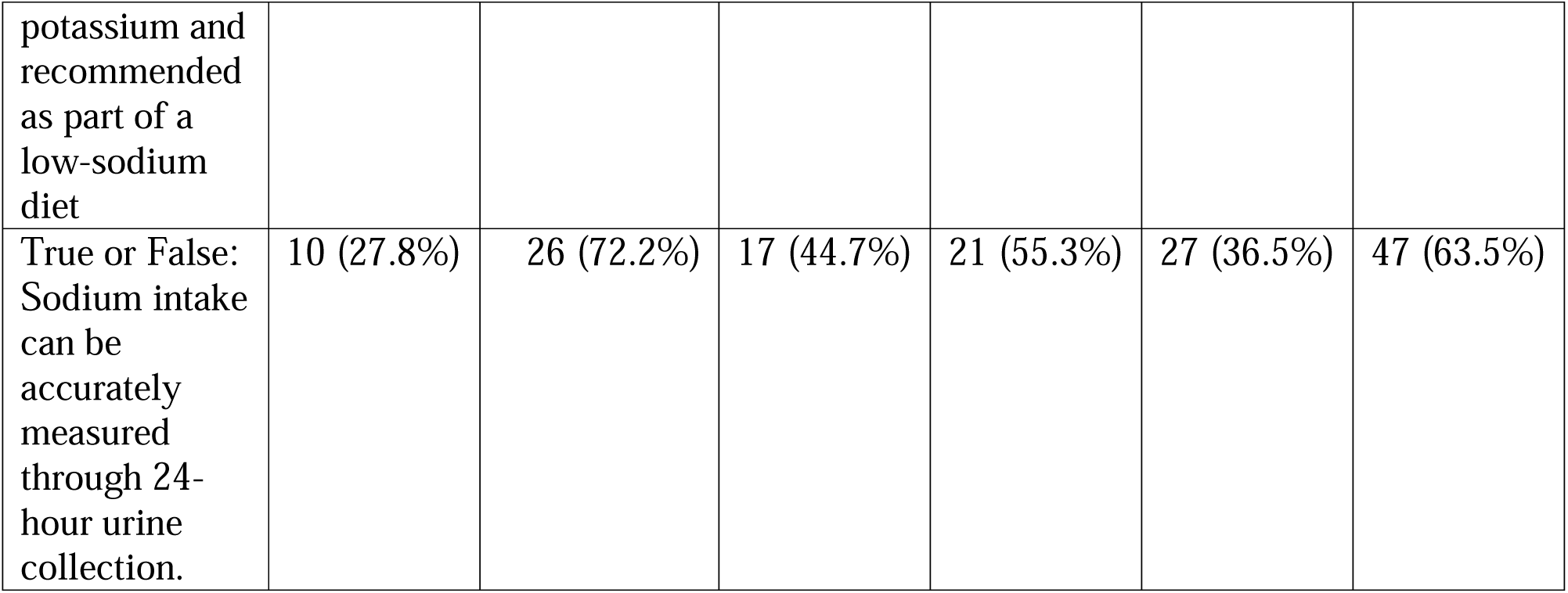
Knowledge Improvement Following the post Workshop in Chennai and Mumbai (n=74)

Although Chennai had a slightly higher average post-workshop score (6.08±1.75) than Mumbai (5.47±1.45), the difference was not statistically significant (p=0.106). This suggests comparable effectiveness of the workshop across both cities, though individual topic performance might differ (Table 4).

**Table 4.** Comparison of Mean Knowledge Scores Between Physicians in Chennai and Mumbai.

| Place | Total Score (Mean±SD) | t-value | p-value |
| --- | --- | --- | --- |
| Chennai (n=36) | 6.08 ±1.75 | 1.639 | 0.106 |
| Mumbai (n=38) | 5.47±1.45 |  |  |

In Delhi, the pre- and post-workshop comparison demonstrated overall improvement in knowledge across several domains. Notably, correct responses on sources of dietary salt, benefits of low-sodium salt substitutes (LSS), and accurate sodium assessment increased post-training. However, some misconceptions persisted, particularly in understanding the sodium-to-potassium ratio and safe use of LSS in patients with comorbidities. This highlights the workshop’s positive impact while indicating areas requiring reinforcement in future sessions (Table 5).

**Table 5.**
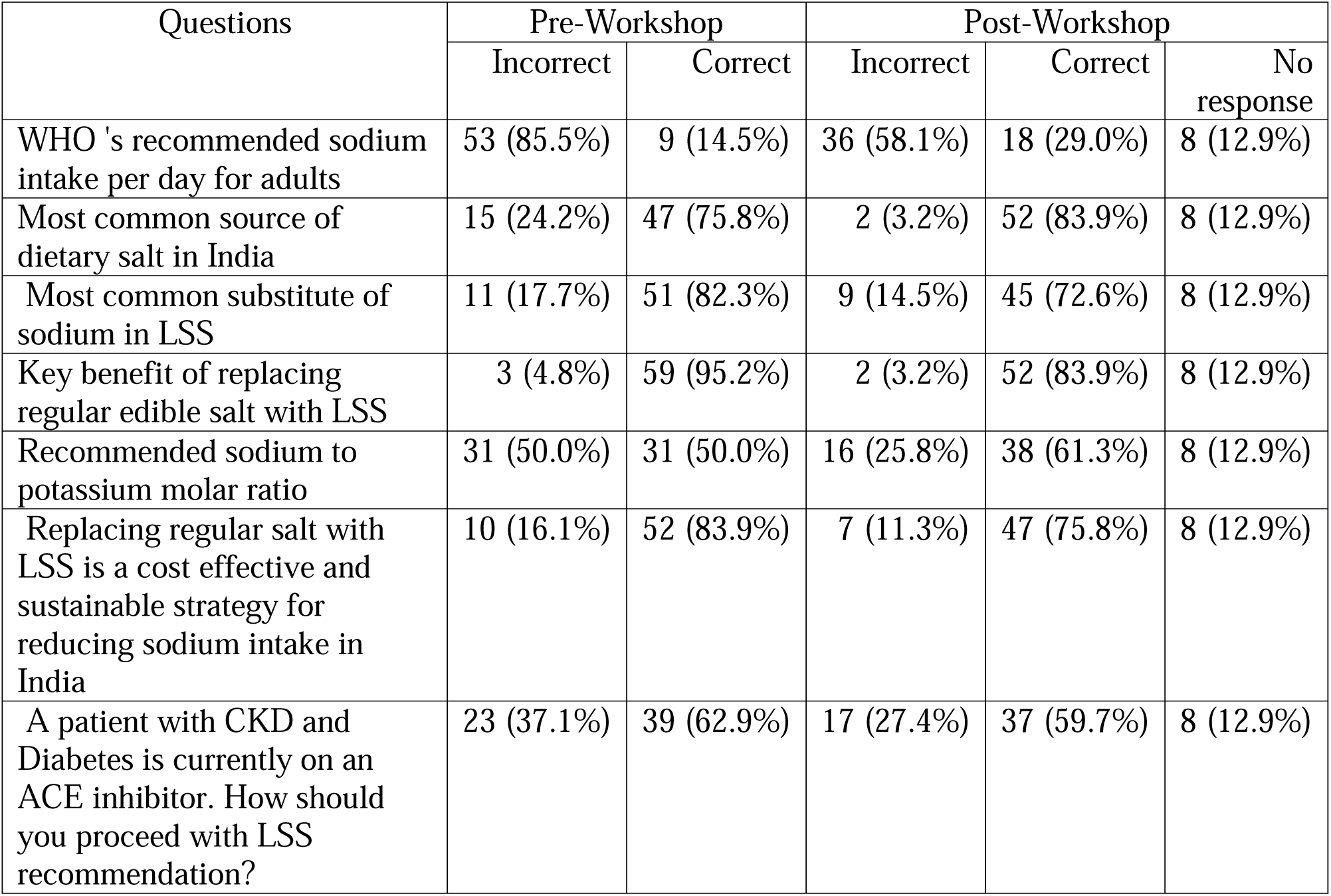
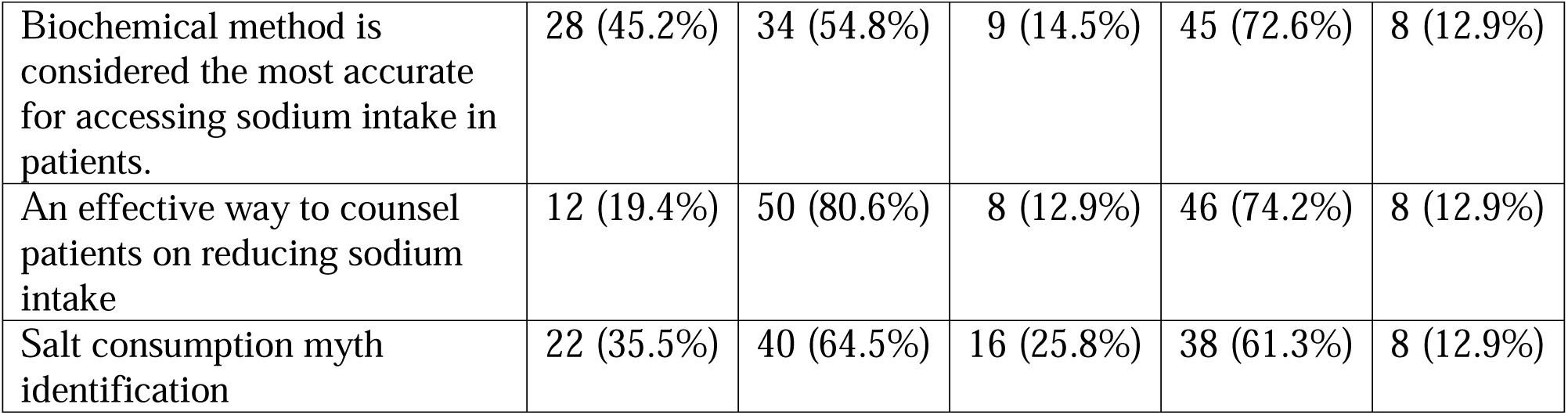
Pre- and Post-Workshop Knowledge Assessment Among Physicians in Delhi (n=62)

| Questions | Pre-Workshop |  | Post-Workshop |  |  |
| --- | --- | --- | --- | --- | --- |
|  | Incorrect | Correct | Incorrect | Correct | No response |
| WHO 's recommended sodium intake per day for adults | 53 (85.5%) | 9 (14.5%) | 36 (58.1%) | 18 (29.0%) | 8 (12.9%) |
| Most common source of dietary salt in India | 15 (24.2%) | 47 (75.8%) | 2 (3.2%) | 52 (83.9%) | 8 (12.9%) |
| Most common substitute of sodium in LSS | 11 (17.7%) | 51 (82.3%) | 9 (14.5%) | 45 (72.6%) | 8 (12.9%) |
| Key benefit of replacing regular edible salt with LSS | 3 (4.8%) | 59 (95.2%) | 2 (3.2%) | 52 (83.9%) | 8 (12.9%) |
| Recommended sodium to potassium molar ratio | 31 (50.0%) | 31 (50.0%) | 16 (25.8%) | 38 (61.3%) | 8 (12.9%) |
| Replacing regular salt with LSS is a cost effective and sustainable strategy for reducing sodium intake in India | 10 (16.1%) | 52 (83.9%) | 7 (11.3%) | 47 (75.8%) | 8 (12.9%) |
| A patient with CKD and Diabetes is currently on an ACE inhibitor. How should you proceed with LSS recommendation? | 23 (37.1%) | 39 (62.9%) | 17 (27.4%) | 37 (59.7%) | 8 (12.9%) |
| Biochemical method is considered the most accurate for accessing sodium intake in patients. | 28 (45.2%) | 34 (54.8%) | 9 (14.5%) | 45 (72.6%) | 8 (12.9%) |
| An effective way to counsel patients on reducing sodium intake | 12 (19.4%) | 50 (80.6%) | 8 (12.9%) | 46 (74.2%) | 8 (12.9%) |
| Salt consumption myth identification | 22 (35.5%) | 40 (64.5%) | 16 (25.8%) | 38 (61.3%) | 8 (12.9%) |

The average score improved slightly from pre- (6.65±1.64) (n=62) to post-workshop (6.74±3.02) (n=54), but the difference was not statistically significant (p=0.831). The wider standard deviation post-workshop may reflect variation in baseline knowledge or engagement during training (Table 6). While the workshop did improve knowledge, future efforts should aim at reducing variability and ensuring consistent understanding across all participants.

**Table 6.** Impact of Workshop on Knowledge Improvement Among Physicians in Delhi (n=62 Pre, n=54 Post)

| Assessment time | Total Score (Mean±SD) | t-value | p-value |
| --- | --- | --- | --- |
| Pre-Workshop | 6.65 ±1.64 | 0.215 | 0.831 |
| Post-Workshop | 6.74 ± 3.02 |  |  |

These findings indicate a need for targeted physician education to address specific gaps in knowledge on salt consumption and its impact on health.

## Discussion

Excessive sodium intake is a major risk factor for hypertension, cardiovascular diseases, and other non-communicable diseases, posing a significant public health challenge globally. While public awareness campaigns and food industry regulations play a role [7], the active involvement of physicians is indispensable for translating scientific evidence into actionable health behaviours.

Building physician capacity through targeted workshops is increasingly recognized as an effective strategy for improving health outcomes by enhancing healthcare professionals’ knowledge, skills, and confidence in various areas. Physicians are also trusted source of information for patients. Globally, healthcare professionals are tasked with delivering high-quality, evidence-based, and patient-centred care amidst an evolving healthcare landscape. To meet these demands, continued professional development through structured, relevant, and practical training is essential [8]. In this context, the current initiative focusing on salt reduction and low-sodium salt substitutes (LSS) addresses a critical area of public health: excessive dietary sodium intake, and aims to enhance physicians’ knowledge and confidence in guiding patients.

Physicians serve as an essential link between scientific guidelines and community health behaviours. Physicians are uniquely positioned to educate patients, diagnose and manage salt-related health issues, and advocate for policy changes. Their direct interaction with patients provides a valuable opportunity to provide personalized dietary advice, monitor progress, and reinforce the importance of salt reduction. Furthermore, physicians can contribute to community-level interventions by participating in health programs, offering expertise, and influencing local health policies. Findings from this multi-city workshop initiative show that while physicians are generally aware of WHO’s recommended salt intake levels, substantial knowledge gaps exist in areas such as hidden salt sources, paediatric salt limits, availability/potential of low sodium salt, and biochemical assessment methods. This aligns with existing evidence suggesting that simple, structured physician-led interventions can lead to modest yet measurable reductions in sodium intake, particularly among high-risk populations such as those with diabetes or renal dysfunction (Oyabu et al., 2021).

Moreover, results from Delhi, where a structured pre- and post-assessment approach was used, demonstrated significant improvement in physician knowledge following the training. These outcomes stress on the value of focused, evidence-based education in enhancing physician competence. Similar observations have been reported in other workshop-based studies, where participants showed improved understanding of disease mechanisms, evidence-based medicine, diagnostic procedures, specific clinical skills, and patient communication skills, leading to improved healthcare outcomes and better patient care [9–13].

Importantly, the WHO and national health authorities emphasize the necessity of such capacity-building efforts, particularly in countries like India, where healthcare worker density remains uneven, with a pronounced urban-rural divide [14,15]

The success of such workshops depends on contextually relevant content, delivery by domain experts, and the use of participatory learning tools. This program incorporated expert-led sessions on salt-related health risks, clinical utility of LSS, public health advocacy, and national dietary guidelines. It also included toolkits for patient education and practice integration, aligning with recommendations for experiential and relatable training formats (Alagboso, 2018).

Digital learning tools could further enhance this initiative. eLearning offers scalability, accessibility, and flexibility for continued physician training, especially in under-resourced or rural settings [16]. While India ((1:834) [17] has surpassed the WHO-recommended doctor-population ratio (1:1000) [14], disparities persist across regions. A hybrid training approach can help ensure more equitable access to medical education and strengthen healthcare delivery in underserved areas.

Therefore, strengthening the knowledge, skills, and resources of physicians regarding salt reduction strategies is paramount. This includes training in evidence-based counseling techniques, understanding of dietary guidelines, and awareness of the broader public health implications of sodium intake. By investing in physician capacity building, nations can significantly enhance their efforts to reduce salt consumption and improve population health outcomes.

## Limitations

This study has some limitations. First, the assessment methods were not uniform across all three cities. In Chennai and Mumbai, baseline awareness was collected through structured response forms and only post-workshop knowledge was assessed, whereas in Delhi, similar pre- and post-training questionnaires were administered. These differences limit the comparability of results across questions, before and after the workshops, and across locations.

Second, some participants in Delhi (n=8, 12.9%) did not respond to the post-test, which may have skewed the mean scores and contributed to the apparent decline (Tables 5 and 6). Furthermore, it is possible that for some participants, the content, structure, or delivery of the workshop was not effective, which may explain the persistence of incorrect answers even after the training. These findings should therefore be interpreted with caution.

Third, the study only measured knowledge immediately after the workshops. There was no follow-up to assess whether physicians retained the information over time or applied it in their clinical practice.

Finally, participation in the assessments was voluntary. This may have introduced response bias, as those who chose not to respond might have differed in their level of knowledge or engagement compared to those who did.

Even with these limitations, the program shows that physician capacity-building workshops are feasible and valuable. As this is a preliminary effort, future programs would benefit from standardized evaluation methods across sites, follow-up assessments to measure retention, and stronger emphasis on long-term application in clinical practice.

## Key Takeaways

- Physicians are pivotal in translating salt reduction guidelines into clinical practice and community health promotion.
- Knowledge gaps persist among physicians in India regarding the health risks of excess salt and the practical use of LSS.
- Structured workshops that combine expert sessions with educational materials and interactive formats can significantly improve physician knowledge and readiness.
- Capacity-building interventions should include pre- and post-evaluation tools to track learning outcomes and guide future content development.
- Capacity-building programs should be evidence-based, tailored to local needs, and supported by digital learning tools for broader and sustained impact.
- Engagement of public health departments and institutional leaders enhances program reach and policy relevance.

## Conclusion

The physician training workshops conducted by Sapiens Health Foundation represent a critical step toward embedding salt reduction advocacy within India’s clinical care system. By equipping physicians with current evidence, practical guidance, and educational tools, this initiative strengthens their capacity to promote healthier dietary behaviours. Expanding this program across more regions, integrating it into medical education curricula, and aligning it with national salt reduction strategies will be essential for sustained public health impact.

## Ethics Approval and Participant Consent

The study consisted of educational workshops and anonymous questionnaire-based assessments conducted among physicians. Participation was voluntary, and no patient data or identifiable personal information were collected. Data were analysed and reported in aggregate form to ensure confidentiality. In accordance with applicable institutional guidelines and national regulations, formal ethical approval and written informed consent were not required. Completion of the questionnaires was considered as implied consent to participate in the assessment and use of anonymized data for evaluation and reporting purposes.

## Data Availability

Individual participant-level data are not publicly available because they form part of an organizational program evaluation dataset. Aggregated data supporting the findings of this study are included within the article. Additional information may be available from the corresponding author upon reasonable request, subject to institutional and confidentiality considerations.

## Author Contributions

RR conceived the comment, provided project administration and resources, contributed to writing, and oversaw supervision and validation. SKS contributed to conceptualization, prepared the original draft, and drafted revisions. MS contributed to supervision, validation, and editing. UBK, VK, BS, EF, KS, SNN, NM, RKK, and ZP provided supervision and validation. SR contributed to project administration and resources. All authors reviewed and approved the final version of the manuscript. All authors had access to the materials used in the development of this comment and take responsibility for its accuracy.

## Data Availability Statement

The study utilised assessment data generated during the physician training workshops. Summary data relevant to the findings are presented within the article tables. Individual-level raw data are not publicly available due to participant confidentiality but may be accessed upon reasonable request to the corresponding author with institutional approval.

## Declaration of Interests

The authors declare that they have no competing interests related to this work.

## Funding

This project was supported by Resolve to Save Lives, which receives funding in part from Bloomberg Philanthropies.

## Acknowledgement

The authors sincerely acknowledge the contributions of Mrs. Lalitha Ravichandran, Trustee of Sapiens Health Foundation, and Mr. Kannan Subramanian and Ms. Sujatha Vijaykumar, Project Coordinators, for their active involvement in the successful completion of the physician capacity-building program on salt reduction. Their support and coordination played a vital role in ensuring the effectiveness and impact of this initiative.

## Notes

### Competing Interest Statement

The authors have declared no competing interest.

